# Oral Anticoagulant Use by Emergency Medical Services Patients in the United States

**DOI:** 10.1101/2023.09.27.23296256

**Authors:** Henry Wang, Mengda Yu, Ching Min Chu, Travis P. Sharkey-Toppen, J. Madison Hyer, Michelle Nassal, Alix Delamare, Joanthan Powell, Lai Wei, Ashish Panchal

## Abstract

**OBJECTIVE:** Oral anticoagulant (OAC) use raises the risk of death in life-threatening conditions such as hemorrhagic stroke, trauma and traumatic brain injury. We sought to describe the national characteristics of Emergency Medical Services patients with a history of OAC use.

**METHODS:** We used prehospital electronic medical record data from 2018-2020 from the ESO Data Collaborative. We included adults (age≥18 years) receiving 911 EMS care. OAC use included warfarin, dabigatran, rivaroxaban and apixaban. We determined the incidence of EMS calls by OAC users as well as their variation by EMS agency. We compared EMS call, patient, and response characteristics between OAC and non-OAC users, including primary impressions and hospital diagnoses.

**RESULTS:** There were 16,244,550 adult 911 EMS events, including 906,575 by OAC users (56 per 1,000 911 events). Compared with non-OAC users, OAC users were more likely to be older (73.6 vs. 56.9 years), white (78.0% vs. 51.4%) and non-Hispanic (84.5% vs. 78.0%). Incidents involving OAC users were more likely at nursing homes, rehabilitation or long-term care facilities (17.0% vs. 9.2%) but less likely to involve trauma (14.7% vs. 18.1%) or cardiac arrest (1.2% vs. 1.4%). Among OAC users, the most common EMS primary clinical impressions were chest pain (7.4%), altered mental status (7.3%), injury (6.5%), abdominal pain (4.3%), and brain injury (2.8%).

**CONCLUSIONS:** In this national series of prehospital events, 1 in 18 adult EMS encounters involved OAC users. These results provide key perspectives on the presentation of the OAC users in EMS care.

## INTRODUCTION

### Background

Life-threatening hemorrhage such as traumatic brain injury, hemorrhagic stroke, gastrointestinal bleeding or vascular or visceral injury from blunt or penetrating trauma, is among the most important time-critical conditions treated by prehospital Emergency Medical Services (EMS).^1,2^ Common EMS interventions for life-threatening bleeding include direct hemorrhage control, tourniquet application, administration of intravenous fluids and vasopressors. Select EMS units are also able to treat hemorrhage with blood products such a red blood cells, whole blood and plasma.^3–5^

### Importance

Oral anticoagulants such as warfarin and direct oral anticoagulants (OAC - dabigatran, rivaroxaban, and apixaban) are widely used in the outpatient setting for treating or preventing a range of medical conditions. For example, OACs may be used patients with to prevent strokes in patients with atrial fibrillation or after prosthetic heart valve replacement. OACs are also commonly used to treat or prevent thromboembolism.^6^ Oral anticoagulant use is increasing internationally.^6–11^ A major pitfall of OACs is that their use increases the risk of serious bleeding, including life-threatening hemorrhage.^6^ Given EMS’s role in the emergent treatment of hemorrhage, knowledge of a patient’s OAC use history is essential, potentially influencing prehospital care and outcomes. Despite the prevalence of OAC use, there are no broad descriptions of OAC use history among patients receiving EMS care.

### Goals of this Investigation

Leveraging a large-scale EMS electronic health record system, we sought to describe the characteristics and course of EMS patients with a history of oral anticoagulant (OAC) use.

## METHODS

### Study Design and Setting

We analyzed data from the ESO Data Collaborative (Austin, TX). This analysis was approved by the Ohio State University Office of Responsible Research Practices.

The ESO Data Collaborative (Austin, TX) is one of the largest out-of-hospital electronic health record systems in the United States. The software product collects clinical information for the EMS encounter, including event characteristics, patient demographics, clinical signs and symptoms, interventions, vital signs, and outcomes. The software contains data validation (forcing) options to require or constrain user entries for select data elements. The current software follows the National EMS Information System (NEMSIS) version 3 standard and the associated data definitions.^12^ More than 2,000 EMS agencies use the ESO electronic health record software system.

The ESO Data Set is a repository of clinical data from EMS agencies that agreed to providing de-identified data for research and benchmarking. The data have been used in a wide range of research studies.^13–16^ In select communities, these EMS records are linked with hospital outcomes data through a proprietary bi-directional Healthcare Data Exchange (HDE) system. Among participating hospitals and EMS agencies, hospital data elements (including ICD-10 diagnosis codes and dispositions) are linked with the prehospital record using HL7 messaging.

### Selection of Participants

We included all adult patients (≥18 years) receiving “911” EMS care with a reported history of oral anticoagulant use. EMS providers reported medication history as obtained during the course of clinical care. OAC used included the reported use of warfarin (Coumadin®), dabigatran (Pradaxa®), rivaroxaban (Xarelto®), and apixaban (Eliquis®). While not part of the primary analysis, we also identified instances of reported injected anticoagulants, including heparin, enoxaparin (Lovenox®), dalteparin (Fragmin®), Fondaparinux (Arixtra®), and Edoxaban (Savaysa®). We included only medications reported as part of the patient’s medical history, not medications administered during EMS care. We did not study other oral medications that may potentiate bleeding such as such as antiplatelet agents (-e.g., aspirin or clopidogrel).

### Outcomes

We focused on clinical variables plausibly associated with or influenced by a history of OAC use. Variables of primary interest included patient characteristics (age, sex, race, ethnicity), incident characteristics (location type, medical vs. trauma, cardiac arrest, census region). We also determined the primary clinical impression reported by EMS providers; we grouped the impressions according to ICD-10 categories as implemented in the NEMSIS v.3 system.^17^ We also identified subcategories within each primary impression major category. For the subset of EMS events linked to hospital data, we identified the primary hospital discharge diagnosis, grouping according to ICD-10 disease category.

### Data Analysis

We determined the number and incidence of adult 911 responses with a reported patient history of OAC use. Using univariate logistic regression, we compared patient, incident and care characteristics between OAC vs non-OAC users. We compared EMS primary clinical impressions between events involving OAC and non-OAC users. Among cases linked to hospital data, we assessed differences in primary discharge diagnoses between OAC and non-OAC users.

## RESULTS

During the study period (2018-2020) there were 16.2 million 911 responses. A history of patient OAC use was reported for 906,575 911 events (55.8 per 1,000 911 events (95% CI 55.7-55.9%)). (Figure 1) The most commonly reported OACs were apixaban (46.0%), warfarin (34.8%), rivaboxarin (17.5%) and dabigatran (2.4%). (Table 1) The use of other injectable anticoagulants was reported for a smaller number of patients. When stratified by EMS agency, the median incidence of OAC use was 54 per 1,000 911 responses (95% CI: 52, 56, min 0, max 500).

**FIGURE 1.**
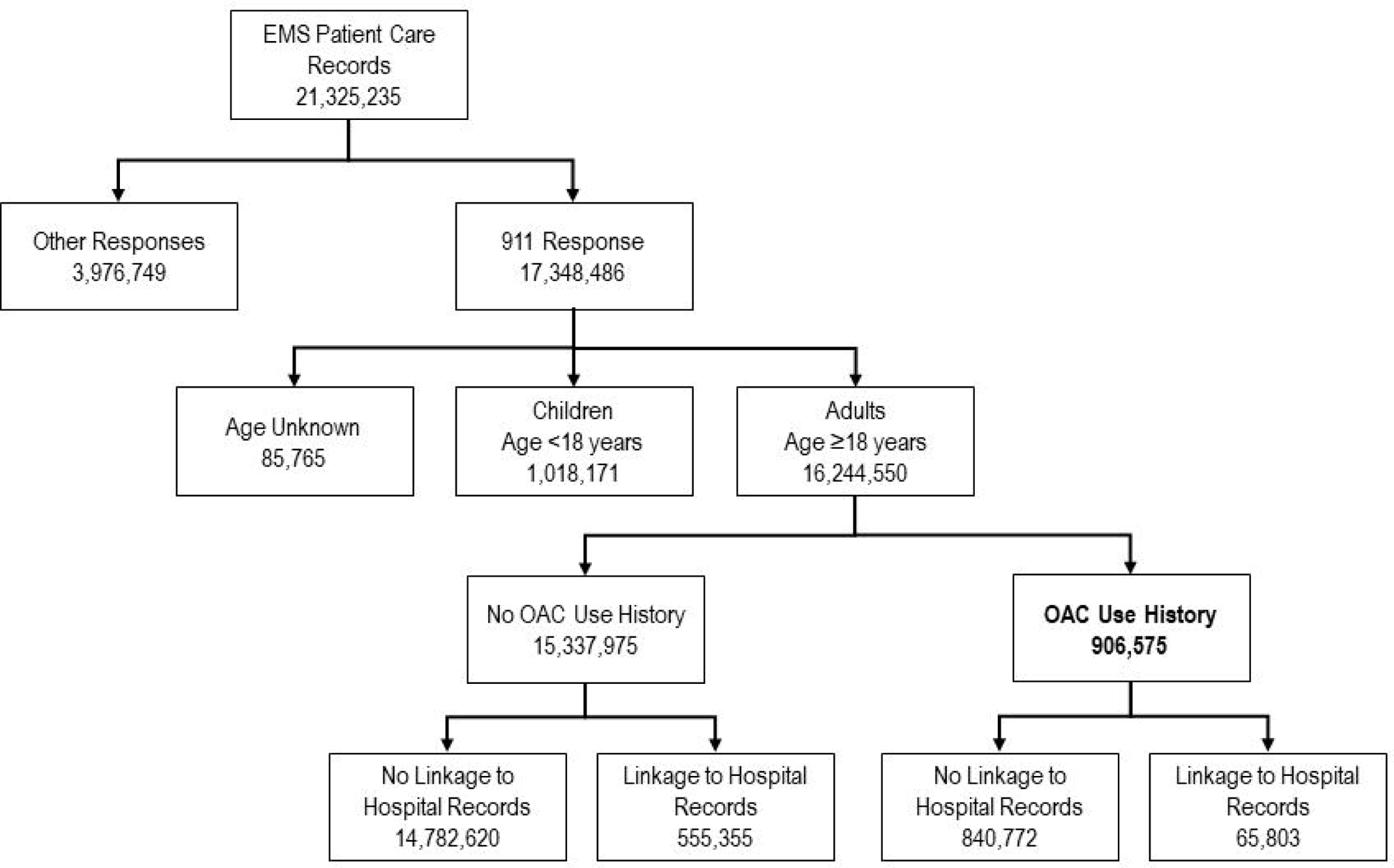
Study population.

**TABLE 1.**
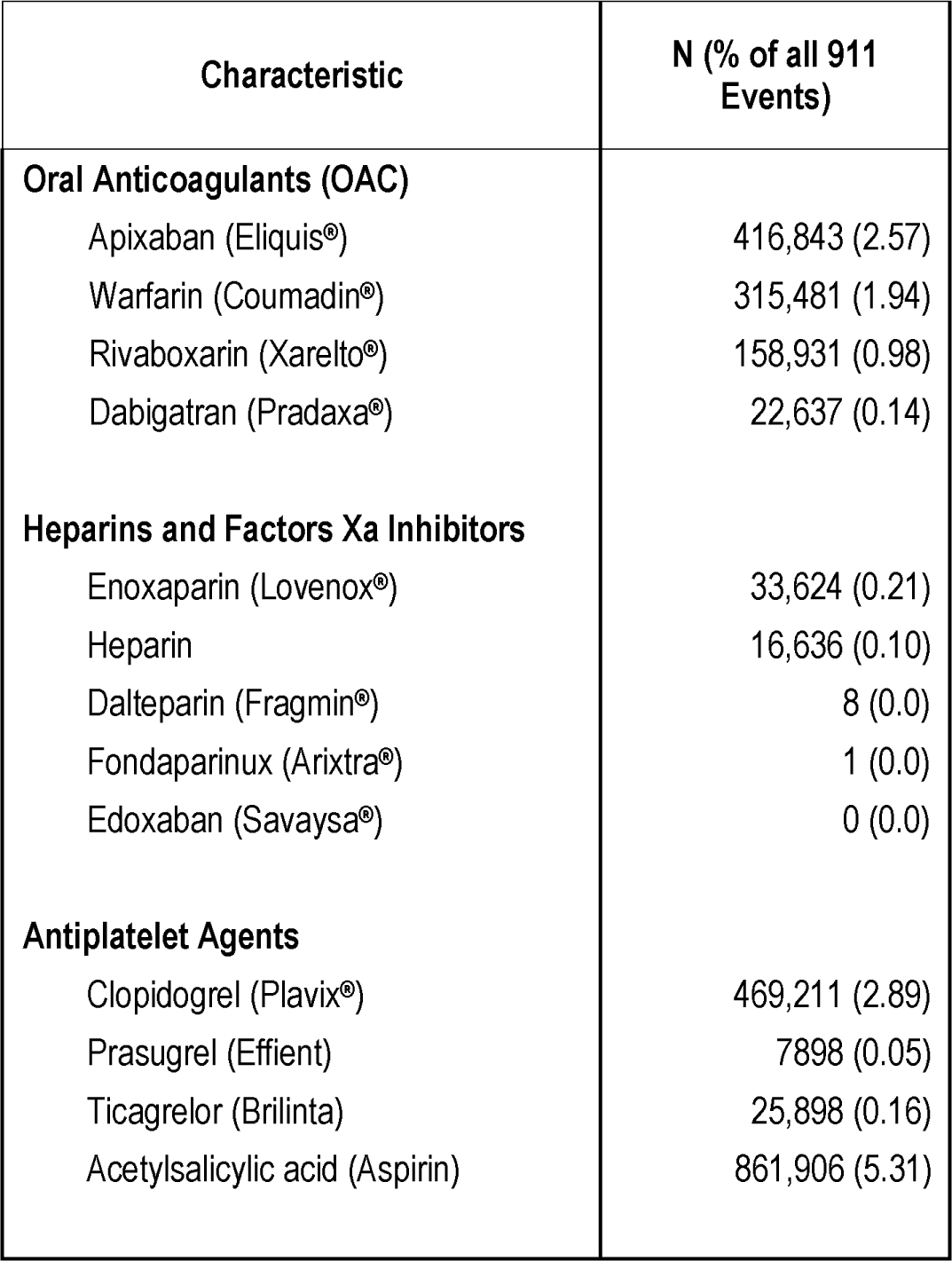
Oral and other anticoagulants reported by EMS patients. Only oral anticoagulants (OAC) were included in the primary analysis.

| Characteristic | N (% of all 911 Events) |
| --- | --- |
| **Oral Anticoagulants (OAC)** |  |
| Apixaban (Eliquis®) | 416,843 (2.57) |
| Warfarin (Coumadin®) | 315,481 (1.94) |
| Rivaboxarin (Xarelto®) | 158,931 (0.98) |
| Dabigatran (Pradaxa®) | 22,637 (0.14) |
| **Heparins and Factors Xa Inhibitors** |  |
| Enoxaparin (Lovenox®) | 33,624 (0.21) |
| Heparin | 16,636 (0.10) |
| Dalteparin (Fragmin®) | 8 (0.0) |
| Fondaparinux (Arixtra®) | 1 (0.0) |
| Edoxaban (Savaysa®) | 0 (0.0) |
| **Antiplatelet Agents** |  |
| Clopidogrel (Plavix®) | 469,211 (2.89) |
| Prasugrel (Effient) | 7898 (0.05) |
| Ticagrelor (Brilinta) | 25,898 (0.16) |
| Acetylsalicylic acid (Aspirin) | 861,906 (5.31) |

Compared with other patients, OAC users were more likely to be older and female. (Table 2) Whites and non-Hispanics were more likely to use OACs. Patient OAC use history was more likely for calls in nursing rehabilitation and long-term care facilities, and less likely for calls in public locations. Compared with medical patients, patients with trauma-associated conditions were less likely to report OAC use. However, OAC use was more common among patients with mixed medical/trauma conditions. OAC use was twice as likely among patients presenting with cardiac arrest. OAC use was least likely among EMS events in the Midwest census region.

**TABLE 2.** Characteristics of EMS patients stratified by history of oral anticoagulant (OAC) use. OAC use defined as use of warfarin (coumadin), dabigatran (Pradaxa), rivaroxaban (Xarelto), and apixaban (Eliquis). Odds ratios determined using univariate logistic regression.

| Characteristic | OAC Use<br>N= 906,575 (5.6%) | No OAC Use<br>N=15,337,975 (94.4%) | Odds Ratio (95% CI)<br>OAC vs no OAC |
| --- | --- | --- | --- |
| **Age (yrs) - Mean (SD)** | 73.60 (14.44) | 56.91 (21.15) | 1.047 (1.047, 1.047) |
| **Age Group (y)** |  |  |  |
| 18-30 | 7,536 (0.8%) | 2,290,499 (14.9%) | Reference |
| 31-40 | 17,482 (1.9%) | 1,800,160 (11.7%) | 2.95 (2.87, 3.03) |
| 41-50 | 36,877 (4.1%) | 1,748,020 (11.4%) | 6.40 (6.24, 6.56) |
| 51-60 | 87,048 (9.6%) | 2,358,358 (15.4%) | 11.20 (10.94, 11.46) |
| 61-70 | 161,712 (17.8%) | 2,490,566 (16.2%) | 19.70 (19.25, 20.16) |
| >70 | 595,920 (65.7%) | 4,650,369 (30.3%) | 38.87 (38.00, 39.77) |
| **Sex** |  |  |  |
| Female | 470,812 (51.9%) | 7,885,062 (51.4%) | Reference |
| Male | 428,773 (47.2%) | 7,253,916 (47.3%) | 0.99 (0.98, 0.99) |
| Unknown | 95 (0.01%) | 3,033 (0.02%) | 0.53 (0.43, 0.65) |
| Missing | 6,895 (0.8%) | 195,964 (1.3%) |  |
| **Race** |  |  |  |
| White | 706,991 (78.0%) | 10,090,534 (65.8%) | Reference |
| American Indian or Alaska Native | 1,019 (0.1%) | 39,668 (0.3%) | 0.37 (0.35, 0.39) |
| Asian | 3,946 (0.4%) | 125,022 (0.8%) | 0.45 (0.44, 0.47) |
| Black or African American | 119,872 (13.2%) | 3,168,567 (20.7%) | 0.540 (0.537, 0.543) |
| Native Hawaiian or Other Pacific Islander | 828 (0.1%) | 20,280 (0.1%) | 0.58 (0.54, 0.63) |
| Other Race | 15,591 (1.7%) | 738,306 (4.8%) | 0.30 (0.30, 0.31) |
| Unknown | 3,723 (0.4%) | 114,747 (0.8%) | 0.46 (0.45, 0.48) |
| Missing | 54,605 (6.0%) | 1,040,851 (6.8%) |  |
| **Ethnicity** |  |  |  |
| Not Hispanic or Latino | 765,913 (84.5%) | 11,964,775 (78.0%) | Reference |
| Hispanic or Latino | 24,423 (2.7%) | 1,056,441 (6.9%) | 0.361 (0.357, 0.366) |
| Missing | 116,239 (12.8%) | 2,316,759 (15.1%) |  |
| **Location Type** |  |  |  |
| Home/Private Residence | 639,900 (70.6%) | 9,033,950 (58.9%) | Reference |
| Doctor' Offices, Clinics and Other Healthcare Facilities | 38,729 (4.3%) | 588,563 (3.8%) | 0.93 (0.92, 0.94) |
| Nursing, Rehabilitation and Long-Term Care Facilities | 154,321 (17.0%) | 1,409,324 (9.2%) | 1.55 (1.54, 1.56) |
| Other | 12,077 (1.3%) | 484,384 (3.2%) | 0.352 (0.346, 0.358) |
| Public Location | 58,105 (6.4%) | 3,652,213 (23.8%) | 0.23 (0.22, 0.23) |
| Missing | 3,570 (0.4%) | 169,414 (1.1%) |  |
| **Medical Trauma** |  |  |  |
| Medical | 727,877 (80.3%) | 11,508,021 (75%) | Reference |
| Medical & Trauma | 27,679 (3.1%) | 379,595 (2.5%) | 1.15 (1.14, 1.17) |
| Trauma | 133,215 (14.7%) | 2,778,524 (18.1%) | 0.76 (0.75, 0.76) |
| Missing | 17,804 (2%) | 671,835 (4.4%) |  |
| **Injured** |  |  |  |
| No | 734,366 (81%) | 120,042,521 (78.5%) | Reference |
| Yes | 160,230 (17.7%) | 3,059,377 (20%) | 0.859 (0.854, 0.864) |
| Missing | 11,979 (1.3%) | 236,077 (1.5%) |  |
| **Cardiac Arrest** |  |  |  |
| No | 5,790 (0.6%) | 234,516 (1.5%) | Reference |
| Yes, After EMS Arrival | 1,373 (0.2%) | 27,547 (0.2%) | 2.02 (1.90, 2.14) |
| Yes, Prior to EMS Arrival | 9381 (1.0%) | 184,315 (1.2%) | 2.06 (1.99, 2.13) |
| Missing | 890,031 (98.2%) | 14,891,597 (97.1%) |  |
| **Region** |  |  |  |
| 1 - Northeast | 80,115 (8.8%) | 854,862 (5.6%) | 1.74 (1.72, 1.75) |
| 2 - Midwest | 193,636 (21.4%) | 3,585,273 (23.4%) | Reference |
| 3 - South | 454,116 (50.1%) | 8,075,588 (52.7%) | 1.04 (1.04, 1.05) |
| 4 - West | 165,095 (18.2%) | 2,447,426 (16%) | 1.25 (1.24, 1.26) |
| Missing | 13,613 (1.5%) | 374,826 (2.4%) |  |

OAC use history was most common among patients with a reported clinical impression involving cardiovascular, infectious or respiratory conditions. (Table 3) Specific primary clinical impressions strongly associated with OAC use history included acute coronary syndromes, arrhythmias, heart failure, gastrointestinal bleeding, hemorrhaging, urinary tract infection, ear, nose and throat problems, pulmonary embolism, and pulmonary hypertension. (Appendix 1) Of note, neurologic conditions including intracranial hemorrhage, vertigo, strokes and transient ischemic attacks and visual disturbances were associated with OAC use. (Appendix 1)

**TABLE 3.** EMS primary clinical impression, stratified by history of oral anticoagulant (OAC) use. Includes 906,575 OAC and 15,337,975 non-OAC patients. Detailed clinical impression subcategories presented in Appendix 1.

| Primary Clinical Impression | OAC Use<br>906 575 (5.6%) | No OAC Use<br>15,337,975 (94.4%) | Odds Ratio (95% CI)<br>OAC vs No OAC |
| --- | --- | --- | --- |
| 1. Cardiovascular | 131,315 (14.5%) | 1,579,867 (10.3%) | 1.475 (1.466, 1.44) |
| 2. Gastrointestinal | 69,888 (7.7%) | 1,283,066 (8.4%) | 0.915 (0.908, 0.922) |
| 3. Endocrine and Electrolyte Abnormalities | 14,885 (1.6%) | 347,088 (2.3%) | 0.72 (0.71, 0.73) |
| 4. Infection | 26,215 (2.9%) | 311,193 (2.0%) | 1.44 (1.42, 1.46) |
| 5. Injury and Environmental Conditions | 136,370 (15.0%) | 2,458,143 (16.0%) | 0.928 (0.922, 0.933) |
| 6. Kidney/Genitourinary | 12,244 (1.4%) | 209,106 (1.4%) | 0.99 (0.97, 1.01) |
| 7. Neonatal | 9 (0%) | 99 (0%) | 1.54 (0.78, 3.05) |
| 8. Neurological | 121,213 (13.4%) | 2,151,632 (14.0%) | 0.946 (0.940, 0.952) |
| 9. Psychiatric | 14,977 (1.7%) | 864,445 (5.6%) | 0.281 (0.277, 0.286) |
| 10. Respiratory and Airway | 116,216 (12.8%) | 1,244,961 (8.1%) | 1.67 (1.65, 1.68) |
| 11. Toxicologic Exposures | 7,209 (0.8%) | 757,298 (4.9%) | 0.154 (0.151, 0.158) |
| 12. Other | 244,563 (27.0%) | 3,781,705 (24.7%) | 1.129 (1.124, 1.134) |
| Missing | 11,471 (1.3%) | 349,372 (2.3%) |  |

Among the 16.2 million adult 911 events, linkage to hospital data was available for 621,158 patients (3.8%). (Table 4) Hospital discharge diagnosis categories most strongly associated with patient OAC use history included D50-D89 (Diseases of the blood and blood-forming organs and certain disorders involving the immune mechanism), I00-I99 (diseases of the circulatory system), and L00-L99 (diseases of the skin and subcutaneous tissue).

**TABLE 4.** Primary hospital ICD-10 discharge diagnoses of EMS patients with a history of oral anticoagulant (OAC) use. Includes 621,158 of 16,244,550 (3.8%) EMS encounters successfully linked with hospital records.

| ICD-10 Group | Description | OAC Use<br>64,344 (10.6%) | No OAC Use<br>541,770 (89.4%) | Odds Ratio<br>(95% CI) OAC<br>vs No OAC |
| --- | --- | --- | --- | --- |
| --- | No diagnosis | 1,186 (1.8%) | 13,465 (2.5%) | 0.74 (0.69, 0.78) |
| A00–B99 | Certain infectious and parasitic diseases | 4,194 (6.5%) | 35,536 (6.6%) | 0.99 (0.96, 1.03) |
| C00–D48 | Neoplasms | 352 (0.6%) | 3,828 (0.7%) | 0.77 (0.69, 0.86) |
| D50–D89 | Diseases of the blood and blood-forming organs and certain disorders involving the immune mechanism | 1,149 (1.8%) | 6,407 (1.2%) | 1.52 (1.43, 1.62) |
| E00–E90 | Endocrine, nutritional and metabolic diseases | 2,574 (4%) | 28,226 (5.2%) | 0.76 (0.73, 0.79) |
| F00–F99 | Mental and behavioral disorders | 508 (0.8%) | 18,109 (3.3%) | 0.23 (0.21, 0.25) |
| G00–G99 | Diseases of the nervous system | 1,573 (2.4%) | 17,245 (3.2%) | 0.76 (0.72, 0.80) |
| H00–H59 | Diseases of the eye and adnexa | 34 (0.1%) | 363 (0.1%) | 0.79 (0.56, 1.12) |
| H60–H95 | Diseases of the ear and mastoid process | 50 (0.1%) | 473 (0.1%) | 0.90 (0.67, 1.20) |
| I00–I99 | Diseases of the circulatory system | 13,062 (20.3%) | 80,695 (14.9%) | 1.46 (1.43, 1.49) |
| J00–J99 | Diseases of the respiratory system | 6,521 (10.1%) | 54,480 (10.1%) | 1.01 (0.98, 1.04) |
| K00–K93 | Diseases of the digestive system | 3,888 (6.0%) | 34,223 (6.3%) | 0.95 (0.92, 0.99) |
| L00–L99 | Diseases of the skin and subcutaneous tissue | 875 (1.4%) | 6,005 (1.1%) | 1.23 (1.15, 1.32) |
| M00–M99 | Diseases of the musculoskeletal system and connective tissue | 1,417 (2.2%) | 10,697 (2.0%) | 1.12 (1.06, 1.18) |
| N00–N99 | Diseases of the genitourinary system | 3,580 (5.6%) | 26,259 (4.9%) | 1.16 (1.12, 1.20) |
| O00–O99 | Pregnancy, childbirth and the puerperium | 9 (0%) | 2,818 (0.5%) | 0.03 (0.01, 0.05) |
| P00–P96 | Certain conditions originating in the perinatal period | 1 (0%) | 22 (0%) | 0.40 (0.06, 2.85) |
| Q00–Q99 | Congenital malformations, deformations and chromosomal abnormalities | 11 (0%) | 121 (0%) | 0.77 (0.41, 1.42) |
| R00–R99 | Symptoms, signs and abnormal clinical and laboratory findings, not elsewhere classified | 14,840 (23.1%) | 115,965 (21.4%) | 1.10 (1.08, 1.12) |
| S00–T98 | Injury, poisoning and certain other consequences of external causes | 5,802 (9.0%) | 61,849 (11.4%) | 0.77 (0.75, 0.79) |
| U00–U99 | Other diagnoses | 820 (1.3%) | 8,909 (1.6%) | 0.77 (0.72, 0.83) |
| V01–Y98 | External causes of morbidity and mortality | 1,229 (1.9%) | 10,363 (1.9%) | 1.00 (0.94, 1.06) |
| Z00–Z99 | Factors influencing health status and contact with health services | 669 (1.0%) | 5,712 (1.1%) | 0.99 (0.91, 1.07) |

## LIMITATIONS

OAC use may have been underestimated. EMS providers likely recorded OAC use based upon clinical data sources such as patient and family reports. It is unclear if reports may have been based upon medication administration or pharmacy records. Medication records may also not be available for patients who were unconscious or exhibited altered mentation. We identified OAC use but not the reasons for their use. We also could not identify the dosages of OACs used by patients. We identified patterns of OAC use and their associations with select conditions; we did not evaluate associations between OAC use and patient outcomes. The latter varies by underlying disease process and requires risk adjustment, which is not standardized for linked EMS-hospital data. While we were able to identify hospital conditions through discharge diagnoses, we were not able to identify the specific pathologic conditions experienced by OAC patients. We did not apply multivariate regression to identify predictors of OAC given the large number of variables and uncertain statistical power. We did not study parenteral anticoagulants nor other agents that may potentiate bleeding such as such as antiplatelet agents.

## DISCUSSION

The history of OAC use is a critical risk factor in life-threatening hemorrhagic and neurologic emergencies. Our study offers important information on the incidence and characteristics of OAC users receiving EMS care. In this national series, 1 of every 18 adult EMS responses involved a patient with history of OAC use. In some EMS agencies the prevalence of OAC use was even higher. As expected, OAC use was common in certain patients with high-risk conditions such as cardiovascular and respiratory diseases, and central neurologic emergencies. OAC use was also strongly associated with EMS responses at nursing, rehabilitation and long-term facilities. These observations highlight the prevalence of OAC use among EMS patients and are particularly important given the increasing prevalence of OAC use internationally.^6–8,10,11^

The knowledge of a patient’s history of OAC use has extremely important implications for EMS care. OAC use may directly amplify the severity and risk of critical conditions, such as hemorrhagic stroke, traumatic brain injury, or gastrointestinal or extremity bleeding.^6,18^ In the setting of acute life-threatening bleeding, knowledge of a patient’s OAC use history may prompt the hospital use of rescue therapies such as blood products (-eg, plasma or red cells), procoagulants (-eg, tranexamic acid, vitamin K, prothrombin complex concentrate) or direct OAC-specific reversal agents (-eg, andexanet, idarucizumab).^6,19^ Because of the potential for rapid deterioration of patients with life-threatening hemorrhage, even the early accurate identification of OAC use history can have important downstream benefits.^20^ The identification of OAC use history may prompt earlier notification to receiving hospitals or transportation to specialty care centers; for example, direct transportation to a trauma center.^20^ Nishijima, et al. found that the addition of OAC use to trauma triaging criteria improved the sensitivity for detection of intracranial hemorrhage and death.^2^ Novel technologies such as mobile stroke units may also be leveraged to accelerate identification or exclusion of intracranial hemorrhage in high-risk OAC users.^21,22^ In certain settings, prehospital administration of procoagulants or OAC-reversal agents may be feasible and appropriate.

Our observations extend upon prior studies. In a retrospective analysis of 2,110 EMS responses for older (age ≥55) persons with head injury, 28% were identified with a history of OAC use.^2^ We found that while many EMS calls for OAC users involved a primary condition of trauma, the majority of events involved non-traumatic emergencies. Prior studies highlight the increasing prevalence of OAC use among older adults and nursing home patients.^9,23,24^ Our study affirms these observations, finding that OAC use was more likely among EMS responses at nursing, rehabilitation and long-term care facilities. Our study spotlights the broader systemwide prevalence of OAC use. The latter approach is important for several reasons. First, we were able to identify OAC use in an EMS population with diverse emergency conditions. Secondly, we able to identify individuals with baseline OAC use - not just those with confirmed hemorrhage.

We were also able to glean insights from the small proportion of patients linked to hospital data. The formation of inferences from linked prehospital/hospital data can be challenging due to the structure of discharge diagnosis coding taxonomies (the International Classification of Diseases) and the myriad medical conditions. We found that focus on key disease subsets was important in identifying hospital conditions associated with patient OAC use. For example, OAC use was not associated with the broad category of neurologic conditions. However, key subset conditions such as intracranial hemorrhage and stroke revealed strong associations with OAC use. The use of linked EMS/hospital data to assess OAC use with hospital outcomes will need careful analytic strategies and taxonomies to identify high OAC-risk conditions.

Our study highlights several important areas for future investigation. Nishijima, et al. found that EMS clinicians often do not accurately identify direct OAC use.^1^ In this context, clinical decision rules may aid in identifying patient and call characteristics at higher risk for OAC use. Our observations highlight several plausible candidate variables such as older age, white race, cardiac arrest, or call locations. Given the wide range of OAC use, additional study is needed to distinguish EMS presentations at high risk for sequelae from hemorrhage. Lastly, geographic mapping of OAC users could be very insightful for system level preparation, identifying hotspots for deploying and positioning resources such as mobile stroke units, procoagulant medications, and novel OAC reversal agents. We observed that EMS calls with OAC users were more common in the Northeast census region, highlight the potential importance of national regional variations in OAC use.

In conclusion, in this national series 1 in 18 adult EMS encounters involved OAC users. While OAC use was common in high-risk populations, presentation with injury was less common. These results offer perspectives on the population of OAC patients receiving EMS care.

## Data Availability

Data are not available from the authors.

## Acknowledgements

The content derived from this Data Set remains the property of ESO Solutions, Inc. ESO is not responsible for any claims arising from works based on the original data, text, tables, or figures. The authors wish to express their appreciation to ESO for its assistance with the data.

## APPENDIX 1

Detailed EMS primary clinical impression, stratified by history of oral anticoagulant (OAC) use. Includes 906, 575 OAC and 15, 337, 975 non-OAC.

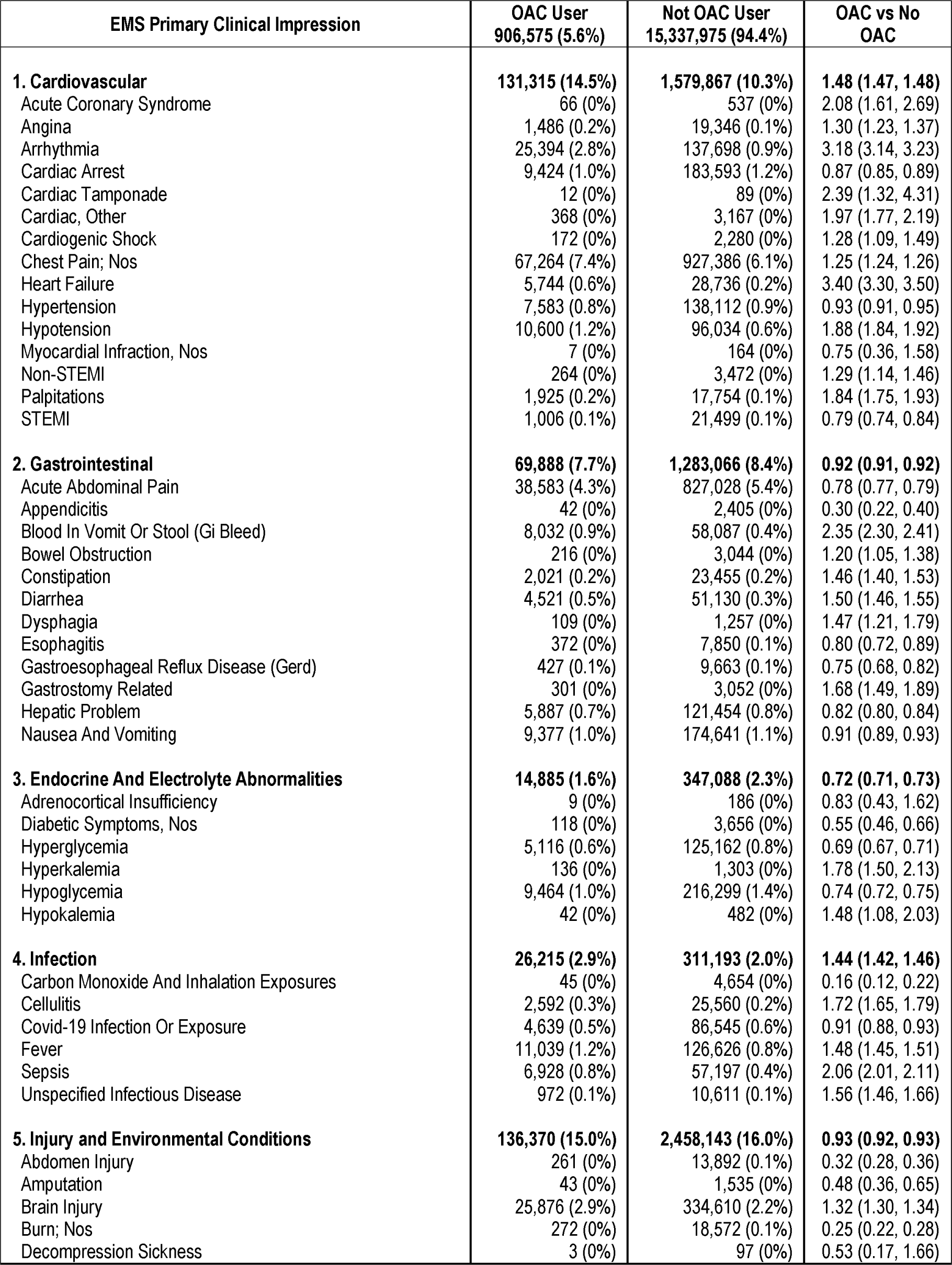
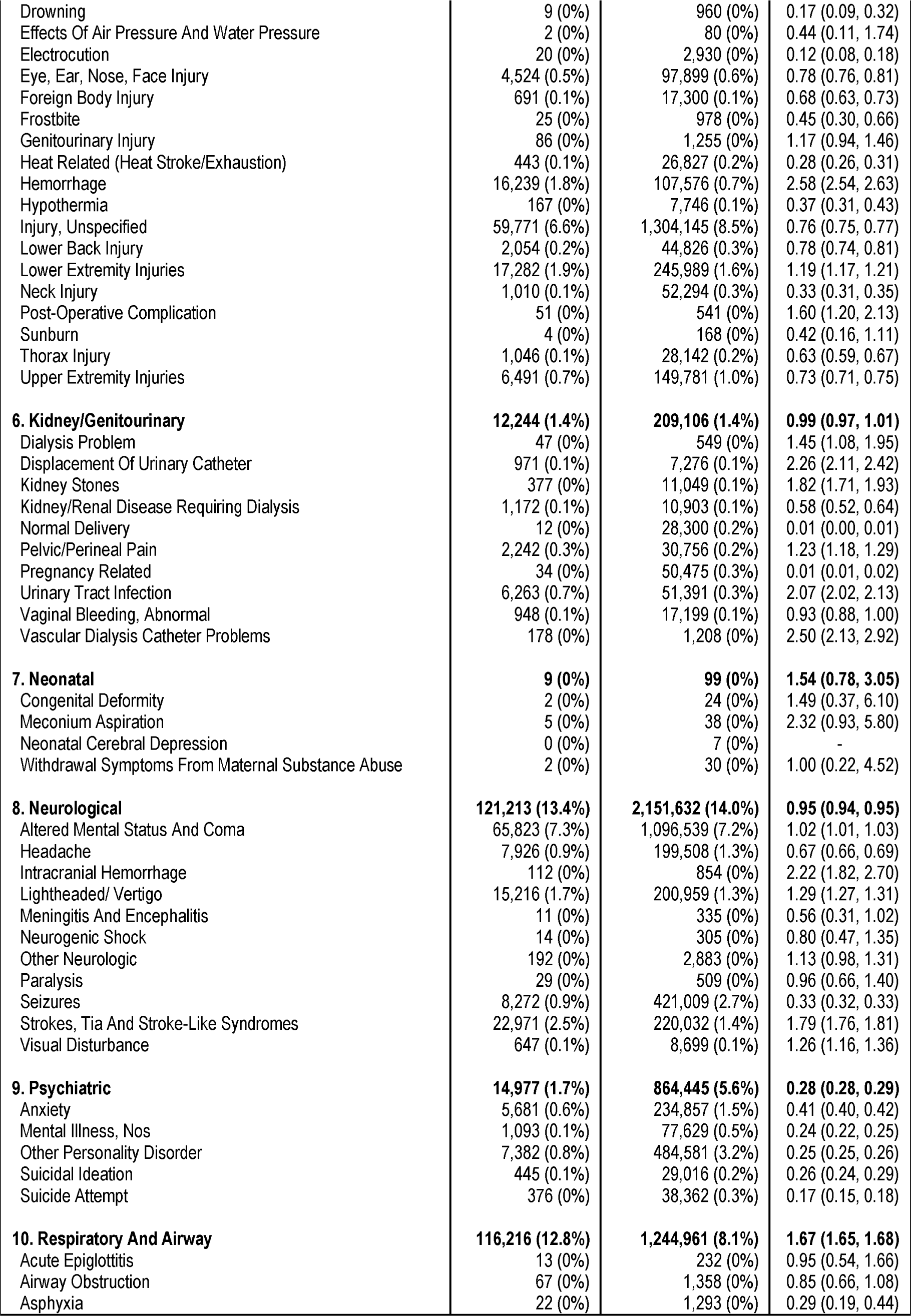
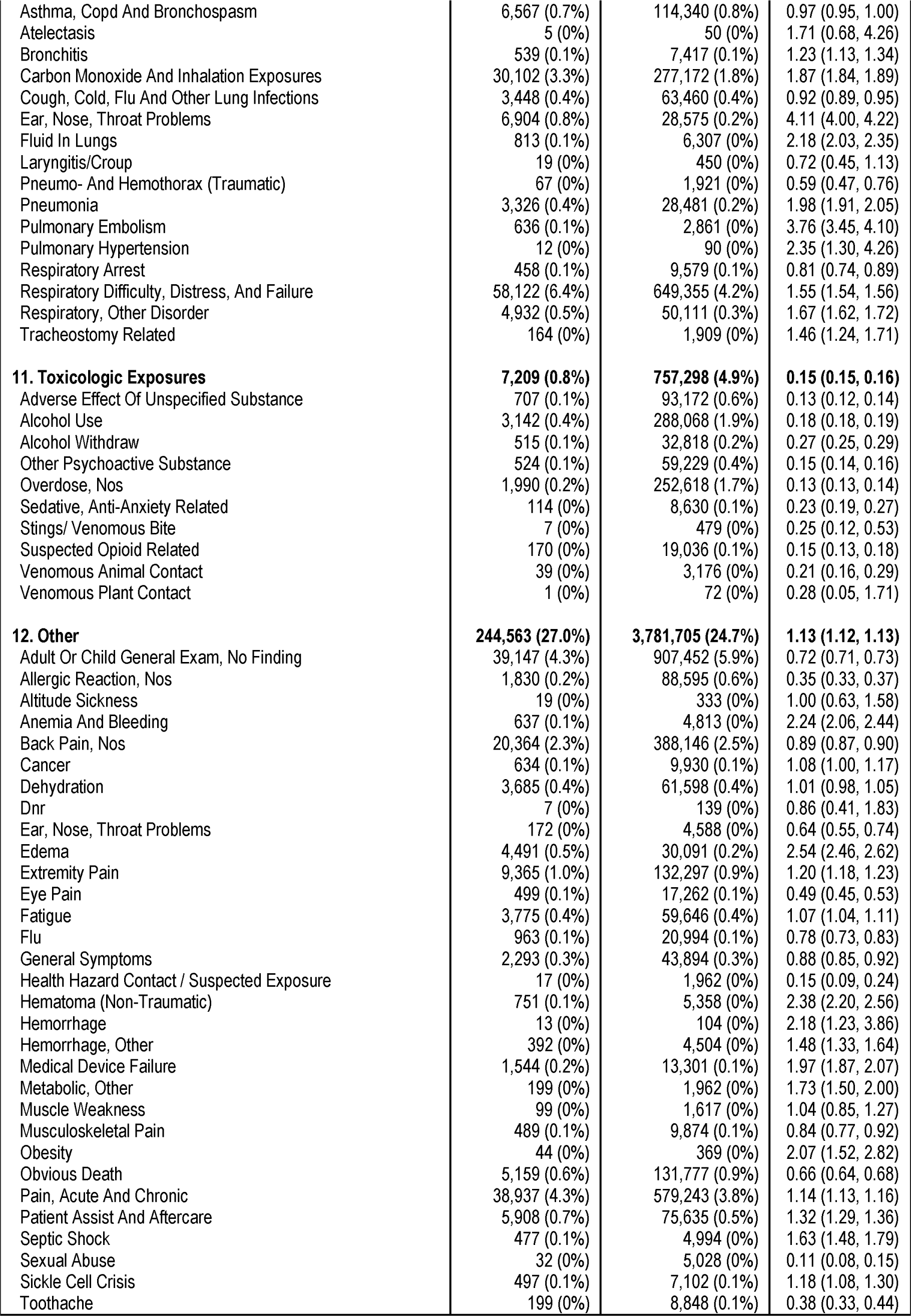
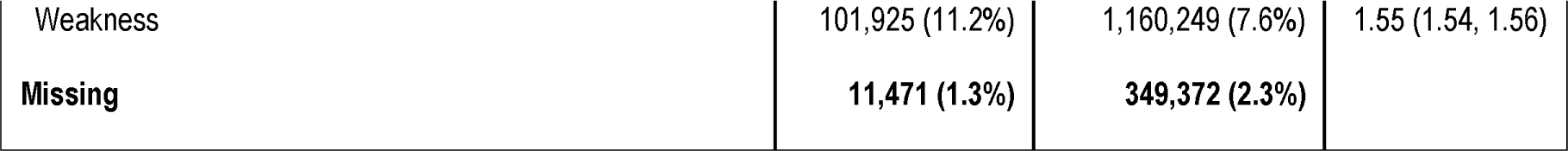

